# Early Versus Late Initiation of Postoperative TPN in Neonates: A Retrospective Study

**DOI:** 10.64898/2025.12.03.25341546

**Authors:** Nao Takami, Akinobu Taniguchi, Kazuto Ueda, Takashi Maeda, Ryosuke Miura, Ryuichi Tanaka, Toshihiko Suzuki, Yukako Muramatsu, Takahisa Tainaka, Chiyoe Shirota, Yoshiaki Sato

## Abstract

**Objectives:** To compare short-term clinical outcomes according to the timing of postoperative total parenteral nutrition (TPN) initiation (early <7 days vs late ≥7 days) in surgical neonates.

**Design, Setting, and Patients:** This single-center retrospective cohort study included neonates admitted to the NICU of Nagoya University Hospital between June 2011 and November 2023. Eligible patients underwent surgery within 30 days of birth and received postoperative TPN. Patients were classified into two groups according to the timing of TPN initiation after surgery: the early TPN group (<7 days; n = 235) and the late TPN group (≥7 days; n = 55).

**Main Outcome Measures:** Infection rate requiring antibiotics.

**Results:** Baseline characteristics were comparable between the groups. No significant difference was observed in the proportion of patients requiring antibiotics within one month after TPN initiation (early TPN group 33.6% vs late TPN group 40.0%, OR =0.76, 95% CI: 0.40-1.46, p = 0.63). Compared with the late TPN group, the early TPN group had a significantly shorter duration of intubation (5.0 days [IQR: 2–18] vs 11 days [IQR: 2.9–34], *p* < 0.01). The incidence of hypoglycemia was significantly lower in the early TPN group (39.9% vs 61.8%, OR = 0.40, 95% CI: 0.21–0.77, p < 0.01).

**Conclusion:** In neonates requiring surgery, delayed initiation of TPN conferred no clinical advantage and was associated with longer intubation and a higher risk of hypoglycemia. Prospective randomized controlled trials are warranted to validate these findings.

• What is already known on this topic –
Early supplementation with adequate nutrition is useful for the development and growth of newborns. However, a previous study suggested that it is better to wait one week before starting TPN in critically ill newborns. There are no reports on whether delaying TPN for one week is beneficial in newborns who are unable to advance enteral nutrition.

• What this study adds –
In newborns who underwent surgery, early initiation of TPN did not increase the infection rate. Moreover, several short-term outcomes were improved in the group that received early TPN.

• How this study might affect research, practice or policy –
This study suggests that early initiation of TPN may provide benefits for postoperative neonates who are at high risk of insufficient enteral intake. These findings highlight the need for prospective randomized controlled trials to further evaluate the role of early TPN initiation in this specific population.

## Background

The optimal timing for initiating total parenteral nutrition (TPN) in neonates remains a critical issue in clinical practice. In particular, neonates requiring surgical intervention often experience delayed recovery of gastrointestinal function and an increased risk of infection, making it challenging to initiate and advance enteral nutrition(1–3). In such cases, the use of TPN becomes essential.

However, the PEPaNIC study, a large randomized controlled trial published in 2016, reported that delaying TPN initiation by one week in critically ill pediatric patients, including neonates, reduced the incidence of infections and shortened the length of ICU stay(4). However, because of these conflicting findings, there is still no clear consensus on the optimal timing of TPN initiation in postoperative neonates, and clinical practices vary.

Following the findings of the PEPaNIC study, our institution revised its protocol in 2019 to delay the initiation of TPN in postoperative neonates, compared with the previous practice of starting TPN within 7 days. This study aimed to evaluate the impact of this policy change on clinical outcomes—such as mortality, infection, hypoglycemia, duration of intubation, and discharge weight—and to re-evaluate the recommended timing for initiating TPN.

## Method

### Study Design and Participants

This was a single-center retrospective observational cohort study conducted in the Neonatal Intensive Care Unit (NICU) of Nagoya University Hospital between June 2011 and November 2023. Clinical data, including gestational age, sex, surgical procedure, date of surgery, and postnatal day of TPN initiation, were collected retrospectively from medical records. Nagoya University Hospital is equipped with a Level IV NICU capable of providing advanced surgical and intensive care for neonates. This study was approved by the ethics committee of Nagoya University Hospital (approval number: 2025-0144).

Eligible participants were neonates who underwent surgical intervention within the first month of life and subsequently received postoperative TPN.

Patients were categorized into two groups based on the timing of TPN initiation: an early TPN group (initiated <7 days after surgery and continued ≥7 days after surgery, before 2019) and a late TPN group (initiated ≥7 days after surgery, from 2019 onward). This classification was determined by a change in institutional protocol for TPN initiation, implemented in 2019 in response to the results of the PEPaNIC trial.

### Outcomes

The primary outcome was the occurrence of infections requiring antibiotic treatment within one month after the initiation of TPN or before discharge. Infection was defined as a condition with clinical signs of inflammation, supported by culture results and the attending physician’s judgment based on symptoms and clinical course.” Secondary outcomes included NICU length of stay, the incidence of hypoglycemia (defined as blood glucose <60 mg/dL) within one month postoperatively or until discharge, the duration of mechanical ventilation within three months postoperatively, and mortality.

Additionally, among preterm infants, anthropometric measurements (body weight, body length, and head circumference) at 36 weeks‘ of corrected gestational age were compared between the groups.

For patients who underwent tracheostomy, the duration of mechanical ventilation was defined as the period during which positive inspiratory pressure (PIP) was required. Cases with incomplete clinical follow-up due to transfer or death were included in the analyses of infection and hypoglycemia when relevant data were available; however, they were excluded from the analyses of NICU length of stay and duration of intubation.

In this study, hypoglycemia was defined as a blood glucose level of <60 mg/dL. Although various thresholds have been proposed in the literature (5), a cutoff of <60 mg/dL - one of the higher values(6) -is used as a clinical indicator for intervention at our institution. This threshold was adopted to facilitate safer evaluation within the context of a retrospective analysis.

### Nutrition Protocol

In our NICU, postoperative neonates receive TPN, consisting of amino acid solution (Pleamin-P®) and lipid emulsion (Intralipos®), both initiated at 0.5 g/kg/day. The dosage is subsequently increased according to the infant’s clinical condition, with a target of approximately 2.0 g/kg/day. TPN is discontinued once enteral nutrition reaches 100 mL/kg/day.

Enteral feeding is initiated within a few days postoperatively, following a comprehensive assessment of abdominal findings and radiographs. Feeding volume is increased daily based on feeding tolerance and abdominal symptoms. Either breast milk or formula is used, and donor milk may be administered for preterm infants. Maternal breast milk is prioritized whenever available.

During TPN administration, metabolic status (such as NH3, bilirubin, triglycerides) is monitored, and the composition and dosage of TPN are adjusted as needed.

### Statistical Analysis

Statistical analyses were performed using EZR (Saitama Medical Center, Jichi Medical University), which is a graphical user interface for R (The R Foundation for Statistical Computing, Vienna, Austria). The distribution of continuous variables was assessed using the Shapiro–Wilk test. For normally distributed variables, Student’s *t*-test was used for group comparisons; otherwise, the Mann–Whitney *U* test was applied. Categorical variables were compared using Fisher’s exact test or the chi-square test, as appropriate. A two-sided p-value <0.05 was considered statistically significant. We used the STROBE reporting checklist when editing(7).

## Results

The study enrollment flowchart is presented in Figure 1. From June 2011 to November 2023, 691 infants underwent surgery at the NICU of Nagoya University Hospital. Among them, 348 infants who received postoperative TPN were identified. Of these, 20 infants who underwent surgery after one month of age were excluded. Among the remaining 328 infants, 273 initiated TPN < 7 days after surgery. Because cases in which TPN was started early and discontinued within a short period were likely to be less severe than those with delayed initiation, 2 infants who were transferred within 7 days after starting TPN (TPN end date unknown) and 33 infants who discontinued TPN within 7 days postoperatively were excluded from the analysis. Ultimately, 238 infants who initiated TPN < 7 days after surgery and continued for ≥7 days were classified as the early TPN group (early TPN group ; n=238). This group was compared with 55 infants whose TPN initiation was ≥7 days after surgery (late TPN group; n=55) (Figure 1). Among them, 45 infants in the early TPN group (19.1%) and 10 infants in the late TPN group (18.2%) were transferred to other hospitals during their treatment.

**Figure 1.**
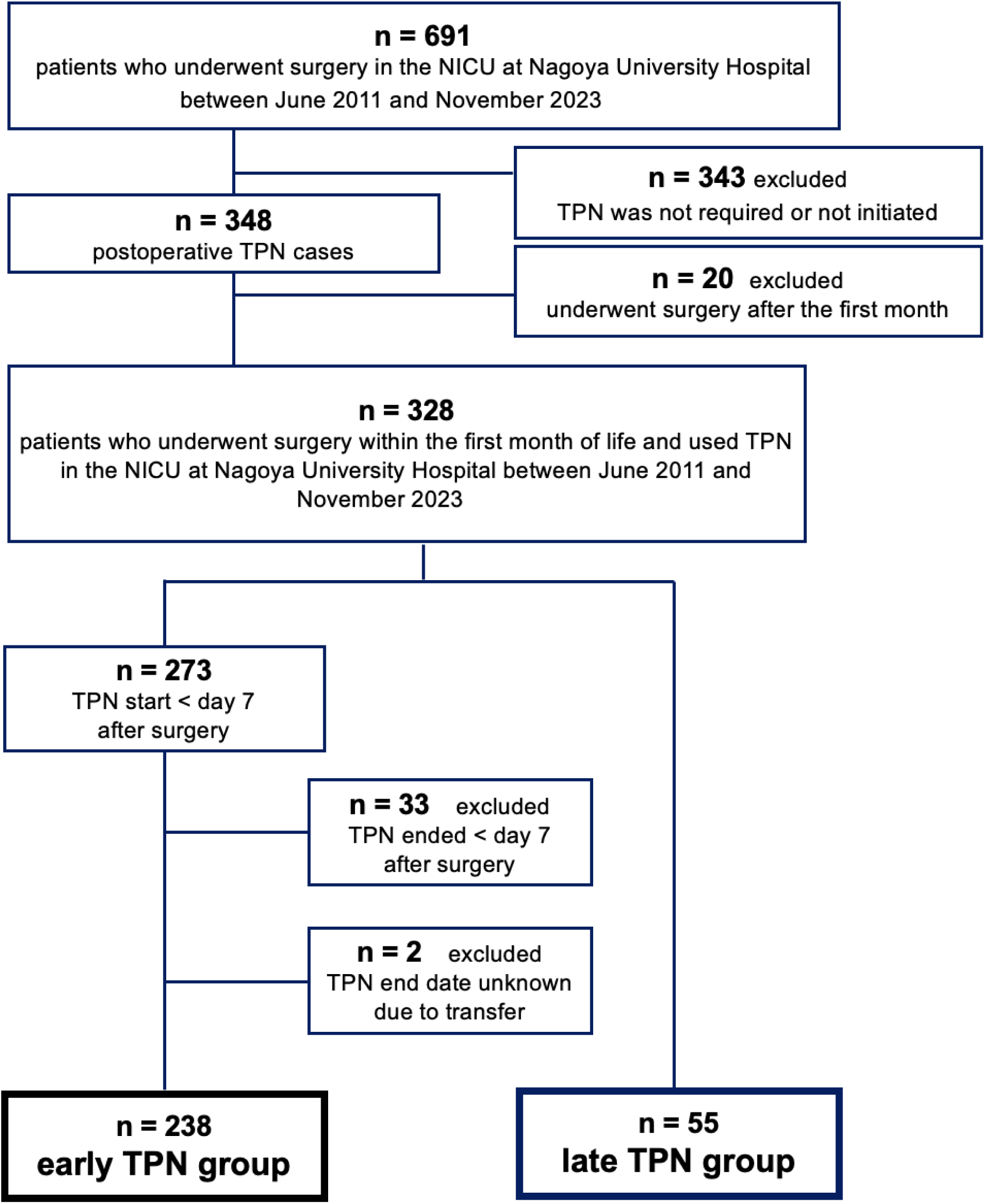
**Study enrollment flowchart**

The median gestational age at birth was 37.3 weeks (interquartile range [IQR], 35.3–38.3) in the early group and 37.1 weeks (IQR, 35.7–38.3) in the late group.

The median birth weight was 2518 g (IQR, 2004–2871) in the early group and 2562 g (IQR, 1884–3014) in the late group. The in-hospital mortality rate was 2.1% in the early group and 1.8% in the late group. The distribution of surgical categories did not differ significantly between the two groups (Table 1).

**Table 1.**
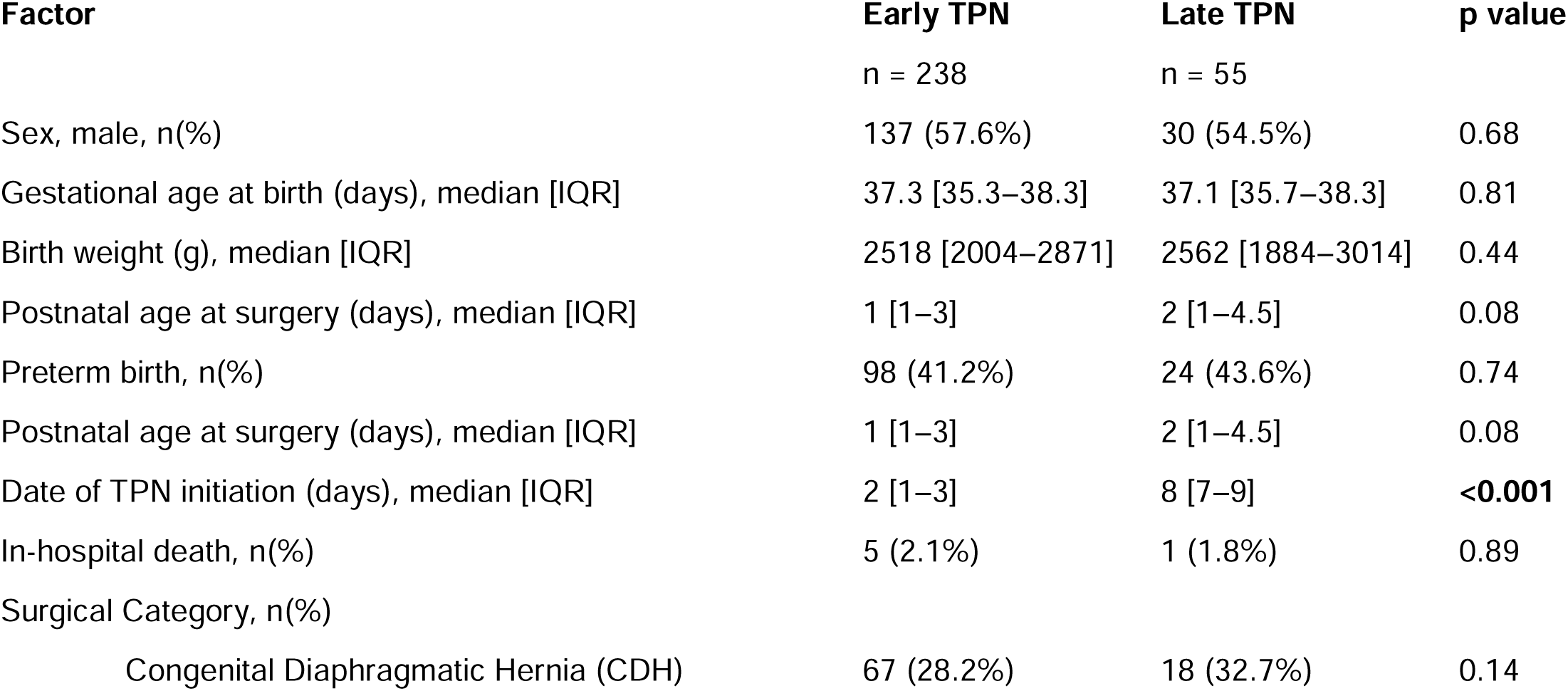
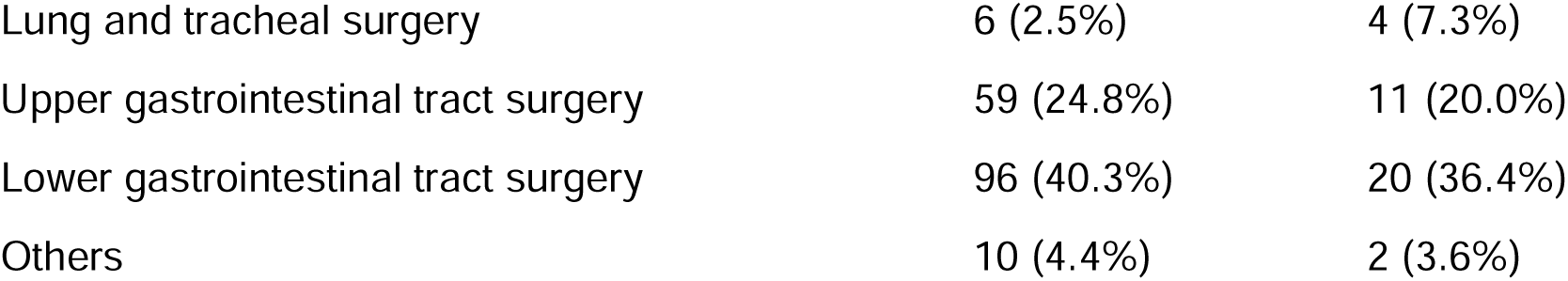
Characteristics of the early and late TPN groups.

| Factor | Early TPN<br>n = 238 | Late TPN<br>n = 55 | p value |
| --- | --- | --- | --- |
| Sex, male, n(%) | 137 (57.6%) | 30 (54.5%) | 0.68 |
| Gestational age at birth (days), median [IQR] | 37.3 [35.3–38.3] | 37.1 [35.7–38.3] | 0.81 |
| Birth weight (g), median [IQR] | 2518 [2004–2871] | 2562 [1884–3014] | 0.44 |
| Postnatal age at surgery (days), median [IQR] | 1 [1–3] | 2 [1–4.5] | 0.08 |
| Preterm birth, n(%) | 98 (41.2%) | 24 (43.6%) | 0.74 |
| Postnatal age at surgery (days), median [IQR] | 1 [1–3] | 2 [1–4.5] | 0.08 |
| Date of TPN initiation (days), median [IQR] | 2 [1–3] | 8 [7–9] | **<0.001** |
| In-hospital death, n(%) | 5 (2.1%) | 1 (1.8%) | 0.89 |
| Surgical Category, n(%) |  |  |  |
| Congenital Diaphragmatic Hernia (CDH) | 67 (28.2%) | 18 (32.7%) | 0.14 |

|  |  |  |
| --- | --- | --- |
| Lung and tracheal surgery | 6 (2.5%) | 4 (7.3%) |
| Upper gastrointestinal tract surgery | 59 (24.8%) | 11 (20.0%) |
| Lower gastrointestinal tract surgery | 96 (40.3%) | 20 (36.4%) |
| Others | 10 (4.4%) | 2 (3.6%) |

No significant difference was observed in the proportion of patients requiring antibiotics within one month of TPN initiation, although the rate was approximately 6% lower in the early group than in the late group (33.6% vs 40.0%; OR =1.3; 95% CI: 0.68-2.45; p = 0.67). In contrast, several clinical benefits were observed in the early TPN group. The median length of NICU stay was not significantly different between the two groups (52 days [IQR: 30–89] vs 64 days [IQR: 44–103], *p* = 0.078). The median duration of intubation was also significantly shorter in the early group (5.0 days [IQR: 2–18] vs 11 days [IQR: 2.9–34], *p* < 0.01). Furthermore, the incidence of hypoglycemia was significantly lower in the early group (39.9% vs 61.8%, OR = 0.40; 95% CI: 0.21–0.77; p < 0.01). (Table 2).

**Table 2.** Comparison of clinical outcomes between the early and late TPN groups.

| Factor | Early TPN | Late TPN | p value |
| --- | --- | --- | --- |
| Infection requiring antibiotics within 1M of TPN initiation, n(%) | 80 (33.6%) | 22 (40.0%) | 0.63 |
| Length of NICU stay (days), median [IQR] * | 52 [30–89] | 64 [44–103] | 0.078 |
| Occurrence of hypoglycemia (BS<60), n(%) | 95 (39.9%) | 34 (61.8%) | **0.003** |
| Days of intubation (days), median [IQR] * | 5.0 [2.0–18.0] | 11.0 [2.9–34] | **0.007** |

\*Death cases are excluded

Among preterm infants with available anthropometric data (80 in the early group and 23 in the late group), anthropometric measurements at 36 weeks of corrected gestational age (body weight, body length, and head circumference) were compared. No significant differences were found between the groups in any of these indices (Table 3).

**Table 3.** Anthropometric measurements at 36 weeks CA.

| Factor | Early TPN | Late TPN | p value |
| --- | --- | --- | --- |
|  | n = 80 | n = 23 |  |
| Body length (cm), median [IQR] | 45.0 [43.4–46.8] | 44.8 [41.3–48.0] | 0.94 |
| Body weight (g), median [IQR] | 32.0 [30.4–33.0] | 31.4 [30.4–32.4] | 0.59 |
| Head circumference (cm), median [IQR] | 2057 [1829–2328] | 1818 [1597–2316] | 0.3 |

## Discussion

In this study, we evaluated the impact of an institutional policy change that delayed the initiation of TPN by one week in neonates who underwent surgery within the first month of life. Delaying TPN initiation did not reduce infection rates, mortality, and NICU stays. Rather, the early TPN group had shorter durations of intubation and a lower risk of hypoglycemia. No significant differences were observed in other major outcomes. These findings suggest that the timing of TPN initiation in postoperative neonates may warrant re-evaluation.

The PEPaNIC study, published in 2016, demonstrated that delaying TPN initiation by one week in critically ill pediatric patients significantly reduced infection rates and ICU length of stay (4). However, the findings of that study differ from ours, potentially due to differences in the study populations. The PEPaNIC study primarily included term infants and did not specifically examine postoperative neonates or preterm infants. The mechanisms underlying the potential benefits of delayed TPN in the PEPaNIC study have not been fully elucidated. Several hypotheses have been proposed, including enhanced ketogenesis (8,9), activation of autophagy(10), and reduced metabolic stress due to amino acid load or impaired protein utilization(11,12).

In contrast, the concept of early initiation of amino acids and lipids after birth has been supported by nutritional guidelines and multiple studies in preterm and very low birth weight infants (13,14). Shen et al. reported that early initiation of TPN was associated with improved growth without an increase in complications (15). Similarly, in a randomized controlled trial by Suganuma et al., early TPN initiation in preterm infants born at 30–33 weeks’ gestation resulted in better weight and length gain compared with infants who received 10% glucose alone(16). Furthermore, Moon et al. reported that early TPN initiation in term and late preterm infants was associated with a significant reduction in postnatal growth restriction(17). Early postnatal growth in preterm infants has also been linked to subsequent neurodevelopmental outcomes(18,19). Notably, a longitudinal MRI study at 7 years of age demonstrated that infants who received higher protein intake during the neonatal period showed more favorable indicators of brain maturation(20).

However, a large observational study by Uthaya et al. reported that although early TPN initiation in extremely preterm infants was associated with improved survival, it was also linked to significantly higher rates of sepsis, bronchopulmonary dysplasia (BPD), retinopathy of prematurity (ROP), and the need for surgical interventions(21). These findings indicate that early TPN should not be universally recommended for all preterm infants and that careful consideration is warranted.

Thus, current evidence regarding the optimal timing of TPN initiation remains inconclusive, with studies supporting both early and delayed initiation. An international survey of PICUs found that many centers did not delay TPN initiation in response to the PEPaNIC study, especially in neonates and malnourished children, because of concerns regarding nutritional restriction (22). Additionally, a survey of NICUs in Australia and New Zealand revealed that while there was variability in the timing of TPN initiation for term and late preterm infants, none of the institutions delayed initiation by more than one week (23).

In adult ICUs, energy deficiency itself has been reported to be associated with increased risk of infections (24). Heidegger et al. reported that early initiation of supplemental TPN in critically ill patients with insufficient enteral nutrition reduced infection rates and shortened the duration of mechanical ventilation in the adult ICU setting (25). Gao et al. also demonstrated that early TPN contributed to a reduction in infections in adult patients undergoing abdominal surgery (26). These findings are consistent with our study and suggest that in postoperative neonates, where enteral feeding is often delayed due to impaired intestinal function, early initiation of TPN may lead to improved short-term outcomes. It is important to note that the patient population in our study—postoperative neonates—differs from that of the PEPaNIC trial, which included critically ill but primarily non-surgical pediatric patients. The transient gastrointestinal dysfunction expected in surgical neonates may explain the differing results. However, the mechanisms underlying the association between TPN timing and clinical outcomes remain unclear. Reynolds et al. reported that, although the number of cases was small, high-dose protein (2.5 g/kg/day) administered postoperatively did not result in clinically apparent protein toxicity in neonates who underwent surgery(27). This suggests that neonates undergoing invasive gastrointestinal surgery with impaired enteral feeding may have higher protein requirements than typical neonates and may therefore be more tolerant of protein loading. Jo et al. reported that newborns who underwent abdominal surgery and achieved the target protein intake exhibited better growth rates(28). In our study, no significant differences in growth parameters were observed between the early and late TPN groups among preterm infants. However, this finding may have been influenced by missing data due to transfers to other institutions. With respect to physical growth and neurodevelopmental outcomes, longer-term follow-up will be required to draw definitive conclusions.

Strengths of this study include a relatively large single-center cohort (n = 293) with standardized care pathways, and a quasi-experimental before–after design anchored to an institutional policy change, which helped mitigate indication bias. However, this study has limitations. First, the retrospective before–after design is susceptible to secular trends and unmeasured confounding. Second, the study population included a wide variety of underlying conditions and surgical procedures, making it difficult to evaluate the impact of TPN timing on specific disease entities. Third, some patients could not be fully followed because of transfer or death, limiting the completeness of outcome assessments. Despite these limitations, our findings suggest the need to reconsider the routine postponement of postoperative TPN in neonates. In particular, for preterm infants and postoperative neonates with delayed advancement of enteral feeding, individualized decisions regarding the timing of TPN initiation and a flexible nutritional strategy may be more appropriate. Future multicenter prospective studies and randomized controlled trials are warranted to optimize the timing of TPN initiation in postoperative neonates and to establish evidence-based nutritional management guidelines.

## Conclusion

In this study, early initiation of TPN in neonates undergoing surgical treatment was associated with clinical benefits, including shorter duration of intubation and a lower incidence of hypoglycemia, without an increased risk of infections or mortality. These findings suggest the need to re-evaluate nutritional strategies that uniformly delay TPN initiation in postoperative neonates. Prospective randomized controlled trials are warranted to establish the optimal timing of TPN initiation in this population.

## Data availability statement

Data are available on reasonable request.

## Patient consent for publication

Not applicable.

## Ethics approval

This study involves human participants and was approved by the Ethics Committee of Nagoya University Hospital (Approval No.2025-0144). The requirement for informed consent was waived owing to the retrospective design of the study, which utilized anonymized data.

## Data Availability

All data produced in the present study are available upon reasonable request to the authors.

## Acknowledgments

This study was presented in part at the Pediatric Academic Societies Meeting 2025, Honolulu, Hawaii. We thank the pediatric surgeons and the medical and nursing staff of the Neonatal Intensive Care Unit at Nagoya University Hospital for their dedicated clinical care and support throughout this study.

## Contributors

NT conceptualised the study, collected data, and drafted the initial manuscript. YS and AT are the corresponding authors and critically reviewed and revised the manuscript for important intellectual content. KU, TM, RM, RT, TS, YM, TT, and CS critically reviewed and revised the manuscript for important intellectual content. All authors approved the final manuscript as submitted and agree to be accountable for all aspects of the work.

